# Role of temperature and humidity in the modulation of the doubling time of COVID-19 cases

**DOI:** 10.1101/2020.03.05.20031872

**Authors:** B Oliveiros, L Caramelo, N C Ferreira, F Caramelo

## Abstract

COVID-19 is having a great impact on public health, mortality and economy worldwide, in spite of the efforts to prevent its epidemy. The SARS-CoV-2 genome is different from that of MERS-CoV and SARS-CoV, although also expected to spread differently according to meteorological conditions. Our main goal is to investigate the role of some meteorological variables on the expansion of this outbreak.

In this study, an exponential model relating the number of accumulated confirmed cases and time was considered. The rate of COVID-19 spread, using as criterion the doubling time of the number of confirmed cases, was used as dependent variable in a linear model that took four independent meteorological variables: temperature, humidity, precipitation and wind speed. Only China cases were considered, to control both cultural aspects and containment policies. Confirmed cases and the 4 meteorological variables were gathered between January 23 and March 1 (39 days) for the 31 provinces of Mainland China. Several periods of time were sampled for each province, obtaining more than one value for the rate of disease progression. Two different periods of time were tested, of 12 and 15 days, along with 3 and 5 different starting points in time, randomly chosen. The median value for each meteorological variable was computed, using the same time period; models with 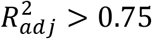 were selected. The rate of progression and doubling time were computed and used to fit a linear regression model. Models were evaluated using *α* = 0.05.

Results indicate that the doubling time correlates positively with temperature and inversely with humidity, suggesting that a decrease in the rate of progression of COVID-19 with the arrival of spring and summer in the north hemisphere. A 20°C increase is expected to delay the doubling time in 1.8 days. Those variables explain 18% of the variation in disease doubling time; the remaining 82% may be related to containment measures, general health policies, population density, transportation or cultural aspects.

## Introduction

The outbreak of pneumonia cases in Wuhan, China during last December led to great efforts to prevent a global epidemic. The alert from China CDC was rapidly transmitted to the World Health Organization,^1-3^ excluding possible causes such as influenza, adenovirus, severe acute respiratory syndrome (SARS-CoV) and Middle-East respiratory syndrome (MERS-CoV).^1,3-4^ The novel coronavirus, named SARS-CoV-2, and its genomic characterization was performed a few days after, permitting to devise a robust test method.^1-5^ Although the genomic characterization revealed some relations both to SARS-CoV and MERS-CoV,^4-6^ the new virus was found to be much more aggressive than those other coronavirus or the seasonal one^4-6^. When human-to-human transmission was proved, on the 20th of January, the onset of the disease (COVID-19) has changed.^2,7-10^ According to the China CDC^11^, the case fatality rate (CFR) was 0.2% at the end of January 2020 and 14.4% of the confirmed cases were considered severe or even critical. In the last week of February, 79441 cases were confirmed worldwide (97% in Mainland China) and the number of deaths was 2620 (95% in Mainland China).^12,13^ The epidemiological curves of COVID-19 in China showed the progression of illness in the outbreak over time from December 8, 2019 up to February 11, 2020,^11^ when there were a total of 72314 confirmed cases as the geo-temporal spread of COVID-19.^11^ At that time, the majority of confirmed cases occurred in the northern hemisphere and until the last week of February 2020 no confirmed cases had been reported in South America or Africa, except for one case in Egipt.^12^ In fact, the first confirmed case in Brazil was reported on February 26, while Algeria and Nigeria reported the first cases respectively on the 25^th^ and 27^th^ of February. The discussion about the COVID-19 epidemic spread in the northern hemisphere, while low temperatures and high humidity are present, and the unknown, although expectable positive impact of spring and summer in sustaining the epidemy, as its spread into the southern hemisphere was not as epidemic, has aroused our question: how do meteorological variables, such as temperature and humidity, modulate COVID-19 duplication time?

Even though there is not yet strong evidence that meteorological conditions may have a role on COVID-19 outbreak or on human transmission, some studies have reported their role in guinea pigs influenza transmission^14^ and enveloped virus survival^15^ in droplets. Some evidence of a faster spreading of diseases in high humidity levels has been reported,^16^ namely for the Legionella disease, although this infection is not caused by a virus. Few papers have been written since mid-February on this topic^17-20^ even though the relationship is not perfectly established and more research is required.

We intend to add value to this discussion by evaluating the meteorological impact on COVID-19 duplication time.

## Material and Methods

The statistical model developed was implemented in two steps: firstly, an exponential model relating the accumulated number of confirmed cases and time was considered. Secondly, the rate of spread was used as dependent variable in a linear model that took as independent variables temperature, humidity, precipitation and wind speed.

Only cases belonging to China were considered as an attempt to control both cultural aspects and policies adopted to contain the virus. Therefore, data from the 31 provinces of Mainland China were gathered from the 23^rd^ January up to the 1^st^ of March, completing 39 days. These data were completed with meteorological variables, comprising temperature, humidity, precipitation and wind speed, collected for the same period, using the Meteostat Application Programming Interface (API).^21^ We searched for meteorological stations containing hourly measurements of these variables for the whole 39-day time period that were closest to the latitude and longitude coordinates that were available in the files that contained the confirmed cases time-series.^13^ These geo-localization coordinates correspond to the geometrical centerpoint of each Chinese province.^22^ When meteorological data from a station was not possible to obtain around that position, another search point was chosen randomly from the set of closest nodes of an XY grid of nodes separated by 0.5 degrees in latitude and longitude and cantered in the originally desired geolocation.

To compute the rate of spread a simple exponential model was assumed, described by:

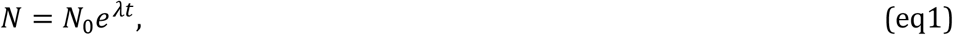

where N_0_ is the number of infected at instant zero, represents the rate of infection or the rate of spread and t is the time. A more natural way of interpreting is by transforming it into the doubling time, T_d_, given by:

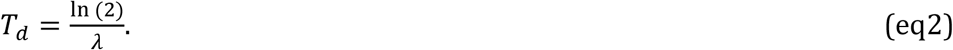

The doubling time is the time needed to duplicate the number of infected subjects. Since the rate of progression changes over time and the exponential model does not hold any longer, we considered mainly the initial days of the time series, selecting several periods of time composed each one of a predetermined number of consecutive days, but with different starting points. The starting point was assumed by randomly choosing the first day of the period. For each province several periods were sampled, allowing to obtain more than one value of the rate of progression (Figure 1).

**Figure 1.**
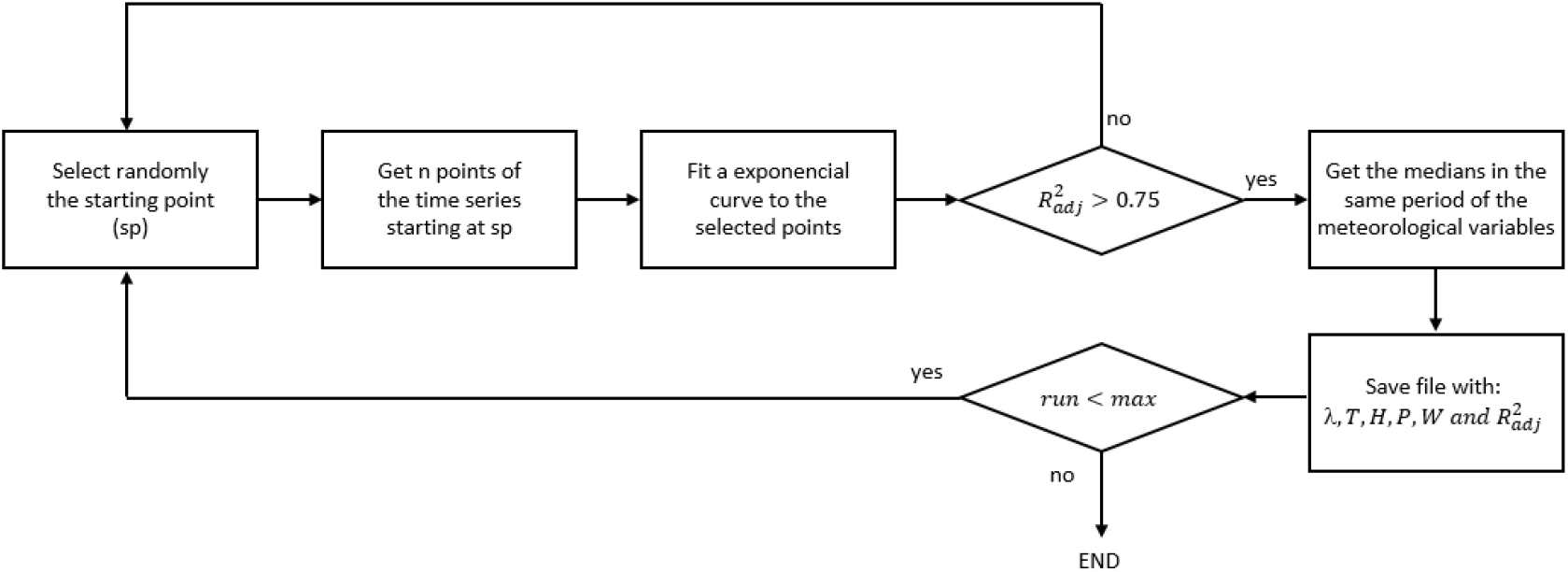
Flowchart of the routine in R language used to compute the statistical models of COVID-19 spreading of new cases

Two different periods of time of 12 and 15 days were tested, along with 3 and 5 different starting points.

For each rate of progression, a corresponding value of each meteorological variable was computed, taking into account the same period of time. We opted to use the median of the meteorological variables (temperature, humidity, precipitation and wind speed) as it is more robust than the mean and tends to better represent the central tendency of the variable. Only models attaining more than 0.75 for the adjusted R square value were selected.

The rate of progression was transformed into the doubling time, T_d_, and recorded along with the median of temperature, humidity, precipitation and wind speed. These values were then used to fit a linear regression model aiming to assess how the meteorology is related to the doubling time.

The exponential models for the rate of progression were compute resorting to the R programming language^23^, whereas the linear regression models were computed using IBM SPSS v25 with an adopted statistical significance level of 0.05.

## Results

The number of confirmed infected cases of COVID-19 were initially analysed by plotting them against time. Figure 2 depicts the curves obtained for the provinces that at the first time point (23^td^ of January) had only one or two cases.

**Figure 2.**
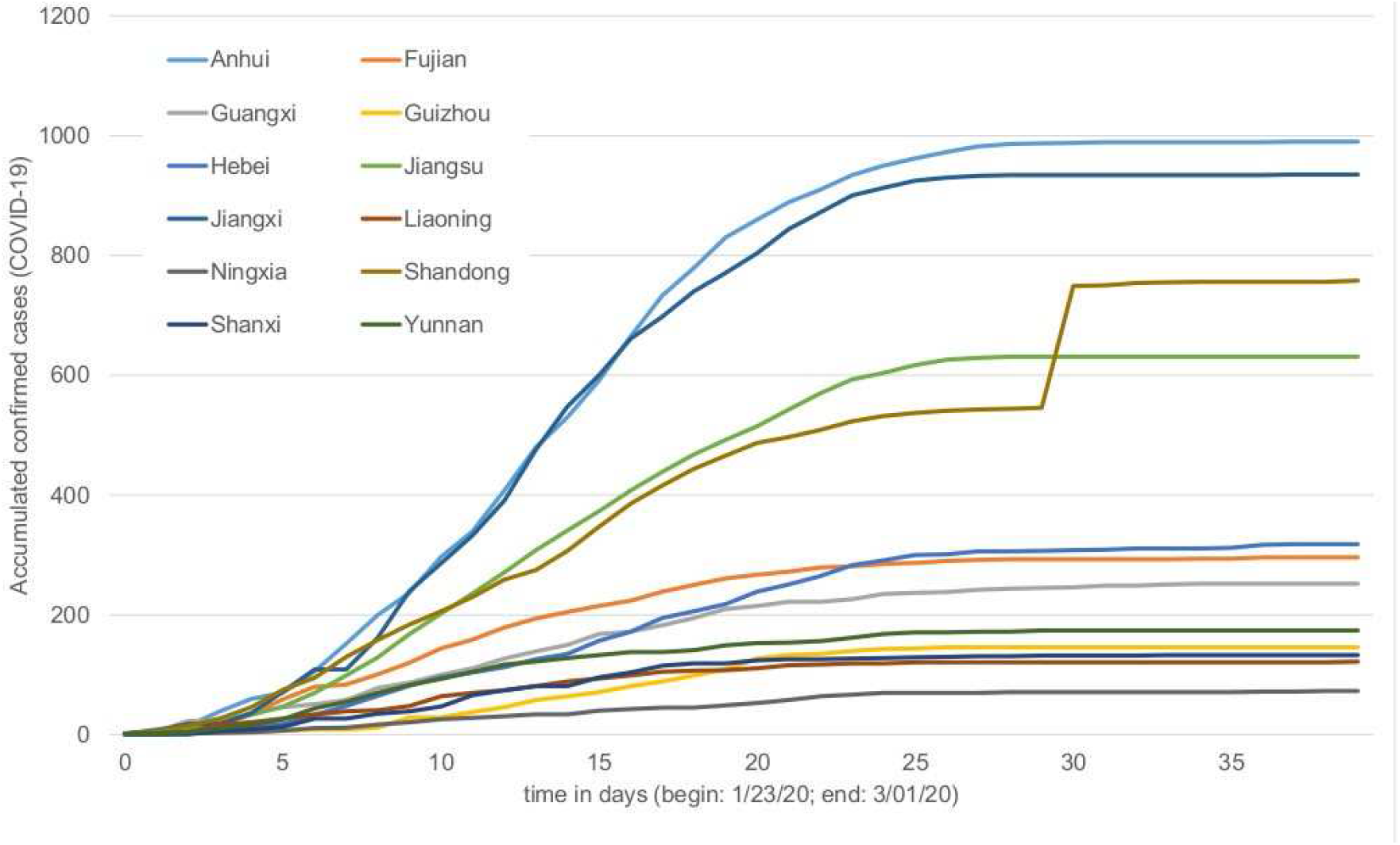
Accumulated confirmed cases of COVID-19 in function of time. Only the provinces that had one or two cases at the beginning of the series are shown.

The analysis of Figure 2 shows that the number of accumulated cases is different between provinces and, in general, its rate decreases over time up to the point where it becomes null. Since the objective of the study was to analyse the rate of spread, we decided to use periods of 12 and 15 days to determine it. The initial 25 days were thought to be the most informative regarding the rate of spread. The fits of temporal evolution of confirmed cases of COVID-19 of the remaining provinces are not shown, but a similar profile can be obtained, leading to the same conclusion. Table 1 shows the statistics - mean, standard deviation, minimum and maximum - of the doubling time, T_d_, for the two ranges of days (12 and 15 days) and the number of sampled periods used for each province.

**Table 1.** Statistics of the doubling time for two different time periods and two repetitions

|  | Period of 12 days | Period of 15 days |
| --- | --- | --- |
| Number of sampled periods = 3 | 3.78 (1.34)<br>1.64/7.05 | 4.53 (1.68)<br>2.04/9.48 |
| Number of sampled periods = 5 | 3.82 (1.27)<br>1.67/7.05 | 4.42 (1.53)<br>1.99/7.76 |
| mean (sd); min/max |  |  |

A two way ANOVA shows statistical differences (F(1, 450)=23.573; p<0.001) between the size, 12 or 15 days of the period employed to compute the doubling time, but no statistical differences (F(1, 450)=0.047; p=0.828) between the number of samples taken from each province. This result is in agreement with the hypothesis that the doubling time changes with time.

The average of the doubling time duplication was determined for each province (Figure 3).

**Figure 3.**
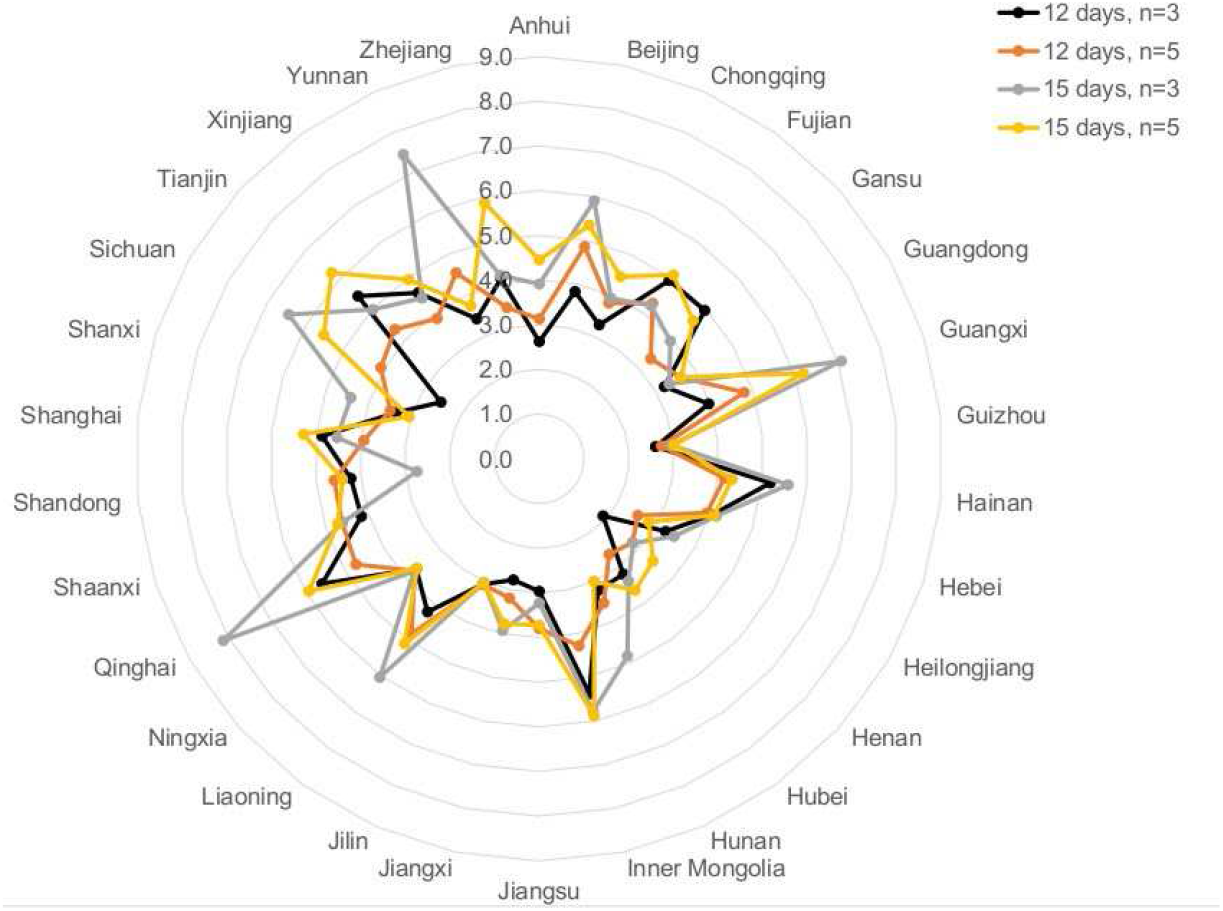
Doubling time for each province considering the different forms of calculation

Taking the values of doubling time T_d_ and the meteorological variables, a linear regression was performed. Table 2 shows, for the 4 conditions studied, the results achieved from the linear regressions, that assumed temperature and humidity as independent variables.

**Table 2.** Results of the linear regressions between doubling time, temperature and humidity.

| Condition | Model significance | $R^2_{adj}$ | Coefficients of regression (B) |
| --- | --- | --- | --- |
| 12 days<br>n=3 | F(2, 81)=10.109 ; p<0.001 | 0.180 | T: B=+0.048 CI95%[0.005; 0.092]; p=0.028<br>H: B=-0.066 CI95%[-0.095; -0.036]; p<0.001 |
| 12 days<br>n=5 | F(2, 140)=8.702 ; p<0.001 | 0.098 | T: B=+0.049 CI95%[0.018; 0.081]; p=0.002<br>H: B=-0.050 CI95%[-0.074; -0.026]; p<0.001 |
| 15 days<br>n=3 | F(2, 83)=6.792 ; p=0.002 | 0.120 | T: B=+0.090 CI95%[0.033; 0.148]; p=0.002<br>H: B=-0.072 CI95%[-0.112; -0.031]; p=0.001 |
| 15 days<br>n=5 | F(2, 138)=6.246 ; p=0.003 | 0.070 | T: B=+0.061 CI95%[0.020; 0.103]; p=0.004<br>H: B=-0.051 CI95%[-0.080; -0.021]; p=0.001 |

T= temperature; H=Humidity

Precipitation and wind speed did not reach statistical significance in any model (data not shown), thus Table 2 only refers results for temperature and humidity. The results obtained for all models are statistically meaningful and despite their variation, it is possible to perceive that the coefficients of regression (B) are not statistically different as their confidence intervals overlap. On the other hand, the amount of variation explained, given by the adjusted R square value, differs between models. The model based on 12 days and 3 sampled periods is able to explain 18% of the variance in the doubling time, which means that temperature and humidity alone may describe 18% of the variation of confirmed COVID-19 infections. More importantly, the signal and value of the coefficients of regressors are of utterly importance to understand how the spread of COVID-19 is expected to be affected by temperature and humidity. According to all models, temperature increases the doubling time, which means that it delays the spread of COVID-19. Humidity, on the contrary, benefits it. The models differ, however, on the amount of contribution: for example, in the best scenario (model: 15 days, n=3) the doubling time is increased by 0.090 days for each Celsius degree increase, and is increased by 0.072 days for each unit decrease of the humidity value.

## Discussion and conclusion

In this work, the way temperature and humidity affect the doubling time of COVID-19 spreading was determined. Results suggest that temperature correlates positively with the doubling time and negatively with humidity. This means that, with spring and summer, the rate of progression of COVID-19 is expected to be slower. Still, these two variables contribute at maximum to 18% of the variation, being the remaining 82% related to other factors such as containment measures, general health policies, population density, transportation, cultural aspects, etc. Besides, the direct impact is also small: for example, if temperature raises 20°C, it is expected that in the best-case scenario the doubling time increases on average 1.8 days.

These results are in agreement with other studies that suggested that the aerosol spread of the influenza virus is both dependent upon relative humidity and temperature, although performed in animal models^14^, and that the virus survival in droplets is higher at high humidity levels with a significant decrease on its infectivity rate at mid-levels of humidity^15^. Additionally, other authors suggest that some diseases spread faster in high humidity levels^16^, reporting an odds ratio for a community-acquired pneumonia case, diagnosed with leggionaire’s disease^a^, 3.1 times higher in high humidity level (above 80%) than when submitted to humidity levels below 50%, at temperatures of 16-27 °C (60°-80°F).

Wei Lo *et al*^17^ recently reported a statistically significant association between absolute humidity and mean temperature on COVID-19 spread among China provinces. Furthermore, they have concluded that transmission and exponential growth of confirmed cases are occurring in China provinces in humidity conditions ranging from cold and dry (Jilin or Heilongjiang) to tropical (Guangxi or Singapore), suggesting that changes in weather, as expected by the arrival of spring and summer, will not necessarily lead to declines in outbreak unless extensive public health interventions are implemented, and that further studies on the effects of meteorology on COVID-19 transmission are needed.

On the other hand, Jin Bu *et al*^18^ reached the conclusion that continuous warm and dry weather is conducive to the survival of the 2019-nCoV and speculate that conditions such as temperature ranging from 13 to 19°C and humidity between 50% and 80% are suitable for the survival and transmission of this new coronavirus. However, their predictions were performed using SARS data and meteorological conditions at that time and, as they report, 2019-nCoV has a high basic case reproduction number (R_0_) lying between 2.2 to 6.7, causing much more infections than SARS.

Moreover, Mao Wang *et al*^19^ have recently submitted a paper supporting that temperature could change the COVID-19 transmission and that there might be an optimal temperature for the viral transmission, suggesting that colder regions in the world should adopt strictest control measures. Yuwen Cai^20^ did not find any correlation between the growth rate of the epidemy and daily mean temperature in either Wuhan or Hunan but found a weak correlation between the mortality in confirmed cases and daily mean temperature both in Wuhan (r = −0.441) and Hubei (r = −0.440), although not adjusted for the use of three makeshift hospitals, which proved to be effective.

The main focus of this work was to assess the relationship between the rate of spread of COVID-19 and some meteorological variables, which determines the type of model adopted. Although the reproduction number, R_0_, is the parameter widely accepted to characterize the velocity of spreading, there are different forms of computing it, which tend to lead to different results^22^. On the other hand, the R_0_ calculation is generally based on assumptions about the epidemic phenomenon such as serial interval distribution^25^ or “the population is closed, that all cases are observed, and use daily case counts only”^26^. For the reasons mentioned, we opted for a simple/naive model that could assimilate the principal aspects of the variation of COVID-19 cases and translate it into a straightforward measurement (T_d_) that could be easily comprehended. Obviously, this model has several drawbacks mostly regarding the optimal period where an exponential growth is verified. We studied two different periods sizes and a random starting point aiming to analyse the impact of this aspect and as an attempt to mitigate its consequences. The doubling time values vary with the period size 26% at maximum (Table 1), which is not neglectable. Even so, this difference only affects slightly the regression coefficients of temperature and humidity, since they do not show statistical differences. Another point of possible bias is the COVID-19 data that do not cover all provinces from the beginning of the outbreak, making it difficult to study homogeneously in all provinces the time period that corresponds to the exponential growth.

Additionally, the meteorological variables in this study were obtained for locations near the centre of provinces, which typically do not correspond to the average location of the population. Measurements that better represent the central tendency of the meteorological variables felt by the general population of a particular region are currently being implemented.

A final remark about the short average doubling time values obtained (3.78 to 4.53 days - Table 1), which should be a motive of concern. For each doubling time, the number of infected doubles, so one month of sustained growth at a conservative pace of 5 days means an increase of the number of infected by a factor of 64.

## Data Availability

Data used in this work is public and is referenced.

## Footnotes

a Caused by bacteria, not virus, but symptoms are similar to flu, with pneumonia as an expected outcome.

